# Perceived and endocrine acute and chronic stress indicators in fibromyalgia syndrome

**DOI:** 10.1101/2024.03.15.24304340

**Authors:** Eva Beiner, Michelle Hermes, Julian Reichert, Kristian Kleinke, Stephanie Vock, Annette Löffler, Leonie Ader, Andrei Sirazitdinov, Sebastian Keil, Tim Schmidt, Anita Schick, Martin Löffler, Micheal Hopp, Christian Ruckes, Jürgen Hesser, Ulrich Reininghaus, Herta Flor, Wolfgang Eich, Hans-Christoph Friederich, Jonas Tesarz, PerPAIN consortium

## Abstract

**Introduction:** Fibromyalgia syndrome (FMS) is a chronic disorder characterized by widespread musculoskeletal pain, fatigue and tenderness and closely associated with high levels of stress. FMS is therefore often considered a stress-related disease.

**Methods:** A comparative study was conducted with 99 individuals diagnosed with FMS and a control group of 50 pain-free individuals. Stress indicators were classified into three categories: perceived stress assessed using the Perceived Stress Scale, and daily average salivary cortisol and hair cortisol concentrations as indicators of acute and chronic stress levels related to the hypothalamic-pituitary-adrenal axis. Analysis of variance and covariance were used to identify group differences and the influence of covariates age, sex, and body mass index. Correlational analyses further elucidated the relationship between stress indicators and clinical symptoms.

**Results:** Participants with FMS reported significantly higher perceived stress levels than controls (*p* < .001, η_p_^2^ = .3), which were positively correlated with symptom burden (*r* = .64, *p* < .001). In contrast, there were no significant differences in the endocrinological stress indicators salivary and hair cortisol between the groups (*p* > .05), nor were these indicators associated with clinical symptoms.

**Conclusion:** The study highlights the central role of perceived stress in FMS, whereas endocrinological indicators did not differentiate FMS from controls. This finding calls for a nuanced approach to clinical assessment and therapeutic interventions tailored to patients with FMS, emphasizing the management of perceived stressors.

## Introduction

Fibromyalgia Syndrome (FMS) has been assumed to be a stress-associated chronic pain disorder [51]. Among the symptom of widespread chronic pain, FMS is characterized by additional somatic symptoms, including fatigue, cognitive malfunction, and sleep disturbances [38]. About 0.2-6.6% of the population is affected by FMS [36]. The disease occurs predominantly in women with a prevalence of 2.4-6.8%, but is also present in men [57]. The pathophysiology of FMS is still under debate, but it is widely assumed that psychobehavioral, social and biological factors play a crucial role in the development [51].

Several lines of evidence strongly support the notion that FMS is a stress-associated disease, with both acute and chronic stress playing important roles [41]. In this context, the interaction of bodily cues, for example, pain, impaired interoception, challenging social contexts and the potential amplification of these factors by acute and chronic stress is emphasized [8; 41]. This multifaceted perspective underscores the critical importance of addressing stress, both acute and chronic, in the assessment and treatment of FMS.

Stress is a latent variable that can be measured through different approaches. Most approaches use either cortisol as an endocrine indicator of stress [28] or questionnaires investigating the experience of perceived stress [27]. Cortisol is used as endocrine stress marker as it rises in response to the exposure to unpleasant stimuli, mental burden, acute demands, or illnesses that induce changes of the body’s homeostasis and activate the hypothalamic-pituitary-adrenal-(HPA) axis. After the activation of the HPA-axis, a cascade of hormones is released in response to the stressor, resulting in elevated cortisol levels to compensate the body’s stress reactivity to sustain homeostasis [4; 28; 40]. Cortisol can be assessed through different methods. It can either be measured through urine, saliva, hair or through blood. During acute stress responses, one can detect elevated cortisol levels in both blood and saliva through single-time-point measurements. However, these measurements prove disadvantageous when aiming to obtain a measure of long-term systemic cortisol production [30; 37]. When evaluating prolonged systemic cortisol production, hair cortisol offers distinct advantages. Due to easy sampling and reduced participant burden, hair cortisol can be easily determined retrospectively for a longer time period. With approximately 1cm growth rate per months, a hair sample of several centimeters can provide the information on cortisol over several months and can serve as an endocrine measure for long-term stress. Consequently, hair cortisol concentration has been suggested as a suitable indicator for assessing chronic stress levels [18; 30; 43].

While an elevated stress experience in patients with FMS has been clinically suggested and documented in the literature [3; 10; 20; 24; 34; 50], there exists considerable inconsistency in the evidence regarding alterations in the endocrine indicators of HPA axis functionality [7; 23; 29; 39; 47]. Hence, there is currently a lack of consensus regarding the relationship between perceived stress levels in people with FMS and the endocrine indicators for acute and chronic stress associated with systemic cortisol production in the HPA axis.

In this study, our primary aim is therefore to identify potential variations in perceived and endocrine indications of stress in individuals with FMS including perceived stress, acute cortisol response and chronic cortisol levels. To explore these variations, we 1.) compared these three indicators of stress between individuals diagnosed with FMS and pain-free controls, 2) investigated the associations between these stress indicators and clinical symptoms, and 3.) examined whether any observed disparities can be attributed to covariates such as age, sex, and body mass index (BMI).

## Methods

This study was part of the PerPAIN study, a large multicenter study, with the goal to phenotype pain patients on a multilevel perspective, to identify subgroups of pain patients according to their pain characteristics and to test whether personalized pain psychotherapy is more feasible compared to non-personalized treatment. The project was funded by the German Federal Ministry of Education and Research. The study protocol was approved by the Ethics Research Committee II of the Faculty of Medicine, University of Heidelberg (2020-579N) and was carried out in compliance with the Helsinki Declaration. For further details on the design of the underlying multicenter study see [5]. This study only used data of the baseline assessment of the PerPAIN study. The preregistrations was uploaded on open science framework (OSF.IO/G5DU7) [6].

### Study design

#### Inclusion criteria

The PerPAIN study recruited a total of 346 participants, from which 320 suited and were screened for eligibility. Finally, 264 individuals participated in the PerPAIN study between April 2020 and August 2023, with 214 individuals suffering from chronic pain and 50 pain-free controls. All participants were assessed by a study physician for the presence of FMS. To be included in this study, participants had to meet the diagnosis of FMS according to the 1990-2016 American College of Rheumatology (ACR) criteria [53–56], be at least 18 years old, must have had symptoms for at least three months, and be able to give informed consent. Participants with secondary pain disorders, acute physical or severe concomitant mental illness, neurological disorders or pregnancy were excluded from this study. To be included in the control group, the participants had to be free of any acute or chronic pain, mentally and physically healthy at the time of the study, which was >assessed by a trained study physician. A flow chart describing the inclusion process can be found in the supplementary material (Appendix A).

### Measurements

#### Stress indicators

To comprehensively map individual stress responses, we collected not only data on the level of perceived stress, but also daily cortisol profiles on two consecutive days to document the short-term activity of the HPA axis as indicator for acute stress, and hair cortisol levels to document the long-term activity of the HPA axis over the previous three months as an indicator for chronic stress.

##### Perceived stress

To measure the level of perceived stress, the German version of the Perceived Stress Scale (PSS) [9] was used. The PSS measures perceived stress through ten items, asking about the participants’ feelings over the last month. Questions include, for example, “*In the last month, how often have you felt nervous and stressed?*”. Answers range from *“0 = Never*” to “*4 = Very Often*”. The sum score was calculated and used for the analysis. The questionnaire is recommended for phenotyping patients for large scale studies [27].

##### Cortisol as acute stress indicator

As a measure of short-term activity of the HPA-axis, salivary cortisol was measured over two consecutive days, adhering to established recommendations for daily average cortisol (DAC) [2; 46]. Participants provided a total of 12 saliva samples—six samples per day. Sampling occurred immediately upon awakening, followed by 15 minutes, 30 minutes, and 60 minutes post-awakening. Additional samples were collected in the afternoon at 3 pm and in the evening at 8 pm, to cover the daily average of cortisol. Saliva samples were frozen (-20°C) until start of analysis. All saliva samples were analyzed in the central laboratory and steroid laboratory of the University Hospital Heidelberg. The standard operating procedures were used, according to the instructions of the manufacturer. They were then centrifuged and analyzed with Liquid Chromatography Mass Spectrometry/Mass Spectrometry (LC-MS/MS) on a Waters Xevo TQ-S System. For the analysis, an average cortisol parameter over both days was calculated [2; 46].

##### Chronic cortisol indicator

Hair cortisol was analyzed to measure the long-term activity of the HPA-axis over a 3-month period. The hair sample was collected from the posterior region of the scalp, with the strand being cut as proximate to the scalp as feasible. All hair samples were analyzed at Dresden University of Technology (TU Dresden). After hair sample extraction, the samples were packed in aluminum foil to be stored dry and dark. The analysis of cortisol was done by using Liquid Chromatography Mass Spectrometry/Mass Spectrometry (LC-MS/MS) method according to Gao et al. [15].

#### Clinical symptom measures

The sociodemographic, clinical, and psychological characteristics of the participants were assessed using an online questionnaire through the REDCap electronic data capture software [19]. Disease duration and tender point count were evaluated by the study physician during the physical examination.

##### Pain Severity and Pain Interference

The severity and interference of the pain experienced were estimated using the corresponding subscales of the German version of the West Haven-Yale Multidimensional Pain Inventory [25]. Both subscales are rated on a 7-point Likert scale and the scores range from 0-6. The final values are derived from calculating the average score. Pain severity was calculated from items covering current pain, average pain over the past week, and the degree of suffering that was induced by the pain. The mean score for pain interference was calculated from items investigating interference with daily life activities such as work, leisure activities or social contacts.

##### Widespread Pain Index (WPI)

The extent of pain was evaluated using the Widespread Pain Index (WPI) [52]. Participants were presented with 19 potential sites on their body and asked to indicate which ones caused pain within the past 7 days. The overall score for the WPI is the total number of identified painful sites, with a range of 0 to 19.

##### Tender points

To examine the tenderness to pressure of individuals, tender points are typically determined. Tender points refer to 18 predetermined areas on the body that are sensitive when pressure is applied. Painful tender points are detected via a tenderness examination adhering to ACR criteria and summed up [52].

##### Somatic Symptom Burden

The Somatic Symptom Burden was evaluated using the 8-item Somatic Symptom Scale (SSS-8) [17]. This scale consists of eight questions that measure the severity of pain in several regions, including abdominal pain, back pain, pain in limbs and joints, headaches, chest pain or shortness of breath, dizziness, fatigue or feelings of depleted energy, and sleep disturbances. Participants rate their symptoms on a 5-point Likert scale, ranging from “*0 - Not at all*” to “*4 - very strongly*”. Sum scores were calculated for analysis, higher scores indicate greater symptom burden.

##### Bodily Distress

To assess the degree of psychological distress caused by somatic disorders, the Somatic Stress Disorder (SSD12) [48] questionnaire was used. With a total of 12 questions, the SSD12 encompasses psychological criteria across three subscales: affect, behavior, and cognition, with four items for each subscale. These items are rated on a 5-point Likert scale from “*0 = never*” to “*4 = very often*”. For the analysis, sum scores were calculated, higher scores indicate higher distress.

##### Polysymptomatic Distress Scale

The polysymptomatic distress scale (PSD) [58] was utilized to assess the impact of fibromyalgia. This scale comprises two components: the Widespread pain index (WPI) as described before and the symptom severity scale (SSS). The SSS measures not only the severity of pain but also assess sleep disorders and cognitive problems. Combining these scales, a total sum score ranging from 0 to 31 is created, with higher scores indicating a more severe symptom burden [58]. Severity categories are as follows: scores between 0-3 indicate no severity, 4-7 indicate mild severity, 8-11 indicate moderate severity, 12-19 indicate high severity, and 20-31 indicate very severe severity. This comprehensive assessment allows healthcare professionals to evaluate and monitor the severity of fibromyalgia symptoms in patients.

##### Pain Duration

Pain Duration was assessed for each individual with FMS in years.

### Statistical Analysis

#### Data preprocessing

Study population, outcomes and main analyses were defined a priori in a statistical analysis plan (OSF.IO/G5DU7). Before the analysis, the data were screened for plausibility of the parameters, out-of-range values, and univariate outliers. This was done by calculating descriptive parameters such as mean and standard deviation, minimum and maximum values, median and the first and third quartile. A missing data analysis was performed for each variable. If missing data occurred, multiple imputation was used to handle the missings in relevant outcomes. This was the case for missing cortisol values. These were substituted by multiple imputation with *m* = 100 using predictive mean matching with the mice package [49]. All data were tested for normal distribution using Shapiro-wilk tests, skew, and kurtosis, and were visually checked for normal distribution using histograms and QQ plots. Furthermore, an outlier analysis was performed, screening for extreme outliers. Any values above or below the cutoff value of the third quartile plus/minus three interquartile distances were defined as extreme outliers and winsorized. Cortisol data were logarithmically transformed to handle extreme outliers and provide normal distribution.

#### Statistical models

For the comparison of each outcome between the two groups (individuals with FMS vs. pain-free controls), an analysis of variance (ANOVA) was performed. Additionally, a two-way analysis of covariance (ANCOVA) was conducted to control for effects of age, sex, and BMI. Further, a pearson correlation analysis was performed to investigate the associations between the outcomes as such, and the associations with clinical characteristics. These included the Widespread Pain Index (WPI), tender points, Somatic Symptom Scale (SSS-8), Somatic Stress Disorder (SSD12), Pain severity, Pain interference from MPI-D and Pain duration. Multiple comparison was handled using Bonferroni-Holm correction [21; 22]. Statistical parameters for ANOVA on imputed data were aggregated accordingly [13; 26]. A p-value < 0.05 will be considered statistically significant. The statistical analysis was performed using R 4.1.2 [42].

## Results

### Descriptive Statistics

A total of 101 individuals with FMS and 50 pain-free controls were eligible for this secondary data analysis. Two individuals with FMS were excluded due to cortisone intake, so a total of 99 individuals with FMS and 50 pain-free controls were included in the analyses. The mean age of the FMS group was *M* = 49.51 (*SD* = 13.08) and *M* = 45.2 (*SD* = 14.62) for the pain-free control group, with no statistically significant difference (*t*(147) = 1.82, *p* = .07, *d* = .31). The FMS group consisted of 87 females (87.9%) and 12 males (12.1%), whereas the control group included 32 females (64%) and 18 males (36%). The BMI was significantly different between the groups (*t* (147) = 3.57, *p* < .001, *d* = .62), with *M* = 27.35 (*SD* = 6.47) for the FMS group, and *M* = 23.82 (*SD* = 3.72) for the pain-free controls. In 31.68% of the individuals with FMS pain existed for more than 20 years, in 22.7% pain existed between 10 and 20 years, in 15.84% pain was present since 5 to 10 years, 23.76% were suffering from pain between one and five years, and in 3.96% pain existed for less than one year Detailed descriptive statistics can be found in Table 1.

**Table 1.** Descriptive Statistics.

| Variable | FMS (n = 99) | Con (n = 50) |
| --- | --- | --- |
| Age, M (SD) | 49.51 (13.08) | 45.2 (14.62) |
| Females, % (n) | 87.9% (87) | 64% (32) |
| Sick leave in days, M (SD) | 22.04 (32.21) | 2.96 (5.11) |
| Working status, % (n) |  |  |
| employed | 77.77% | 86% |
| unemployed | 5.05% | 0% |
| retired | 17.17% | 14% |
| BMI (kg/m<sup>2</sup>), M (SD) | 27.35 (6.47) | 23.82 (3.72) |
| Spatial extent of pain, M (SD) | 11.82 (3.04) | 0.64 (1.06) |
| Somatic symptom burden, M (SD) | 16.97 (5.54) | 3.08 (3.67) |
| Bodily Distress, M (SD) | 25.76 (9.65) | 3.1 (5.06) |
| Tender Points, M (SD) | 11.61 (4.65) | 0.34 (0.89) |
| Pain Severity, M (SD) | 3.56 (0.99) | 0.30 (0.61) |
| Pain Interference, M (SD) | 3.42 (1.25) | 0.30 (0.76) |
| Polysymptomatic Distress Scale, M (SD) | 19.47 (4.07) | 2.06 (2.22) |
| Pain Duration (years), M (SD) | 16.08 (13.2) |  |
| Perceived Stress, M (SD) | 20.81 (6.65) | 11.62 (6.72) |
| Salivary Cortisol in ng/ml | 2.84 (1.69) | 3.05 (1.91) |
| Hair Cortisol in pg/mg | 8.81 (13.53) | 11.55 (23.26) |

*Note.* M = mean, SD = Standard deviation, n = Sample size, BMI = Body Mass Index, Spatial extent of pain = Widespread Pain Index, Somatic symptom burden = SSS-8, Bodily Distress = Somatic Symptom Scale (SSD-12)

### Group differences

For the analysis of group differences of the three stress dimensions PSS, salivary cortisol and hair cortisol, an analysis of variance (ANOVA) was performed in a first step. The results showed that there is a significant difference in subjectively perceived stress between individuals with FMS (*M* = 20.81, *SD* = 6.64) and pain-free controls (*M* = 11.62, *SD* = 6.72), with *F*(1,147) = 63.03, *p* < .001, η_p_^2^ = .30. Individuals with FMS demonstrate higher perceived stress compared to pain-free controls. For salivary cortisol, no significant difference between the groups was found with *M* = 0.49 (*SD* = 0.06) for the FMS group and *M* = 0.57 (*SD* = 0.08) for the pain-free controls (*F*(1,63367) = .52, *p* =

.470 , η_p_^2^ = .004). The same applied to group differences on hair cortisol *F*(1,8427) = .28 , *p* = .596, η_p_^2^ = .003, with a mean of *M* = 1.60 (*SD* = .11) for individuals with FMS and *M* = 1.53 (*SD* = .20) for pain-free controls. Group differences are shown in Fig. 1.

**Fig. 1.**
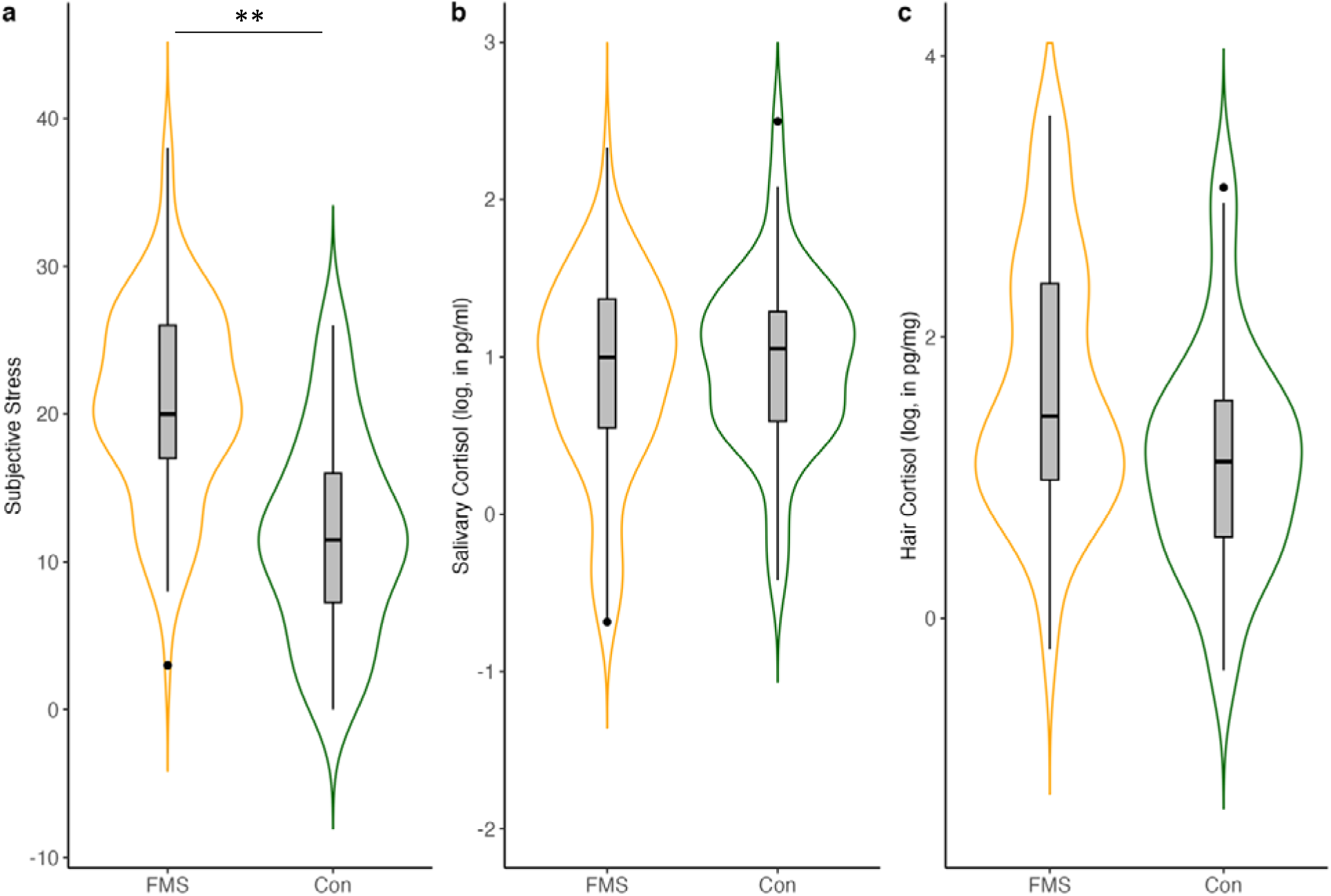
Violin Plots showing group differences of stress indicators a) perceived stress, b) log-transformed salivary cortisol and c) log-transformed hair cortisol. The boxplots represent the distribution of the stress dimensions, with the mean (straight middle line) and the upper and lower interquartile ranges (1.5-fold). Points outside the boxplots represent outlier. FMS = Fibromyalgia Syndrome, Con= Controls. ** p < .001

To explore whether the observed differences might be explained by covariates, a two-factor ANCOVA with the covariates sex, age, and BMI was performed. The total model for perceived stress showed significant results (*F*(1,143) = 23.34, *p* < .001, η_p_^2^ = 0.14), indicating significant group differences. The covariates sex and BMI did not have a significant influence on the model, but age demonstrated a significant influence with *F*(1,143) = 5.57, *p* < .05, η_p_^2^ = 0.046, for perceived stress with greater values for the FMS group. For salivary cortisol and hair cortisol, the covariates did not demonstrate any significant effects on the group differences.

### Associations of stress dimensions and clinical symptoms

For the examination of the associations among the stress indicators and clinical symptoms, a correlation analysis was performed. Significant associations were found between perceived stress and somatic symptom burden (*r* = .64), bodily distress (*r* = .68), pain extent (*r* = .51), number of tender points (*r* = .44), pain severity (*r* = .63), pain interference (*r* = .68) and pain duration (*r* = .35), with a *p* < .001. Overall, higher perceived stress is associated with greater symptom severity. Additionally, we found positive associations between the Fibromyalgianess score (PSD) and perceived stress (*r* = .58, *p* < .001). No significant association were found between salivary and hair cortisol and any of the clinical outcomes. Furthermore, no significant relationship between the stress dimensions themselves were found. Detailed results for the correlation analysis are available in Table 2 and Fig. 2.

**Fig. 2.**
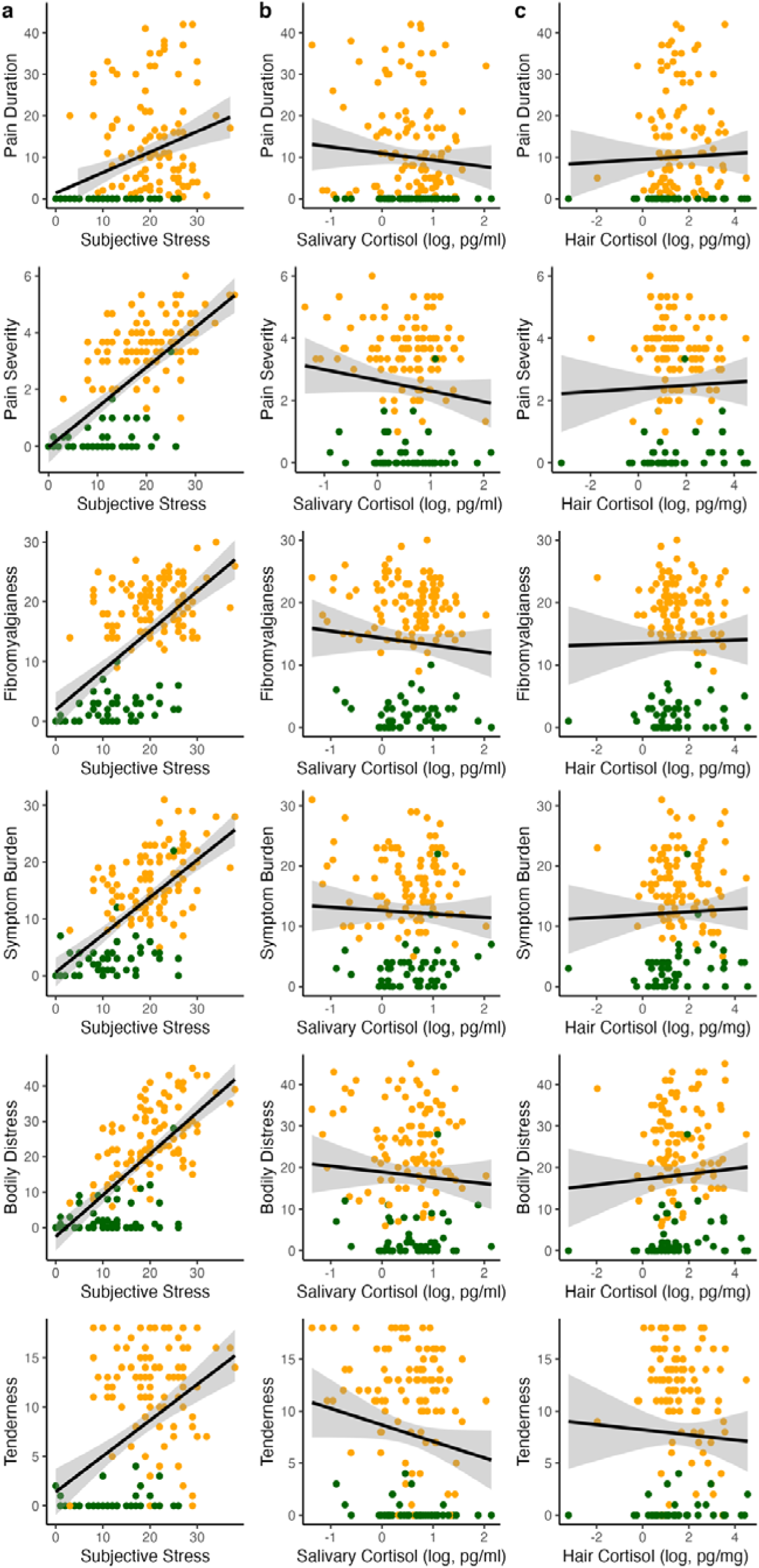
Correlations between the three stress indicators (a) perceived stress, (b) log salivary cortisol and (c) log hair cortisol with clinical outcomes

**Table 2.** Correlation coefficients of stress indicators for the complete sample N = 149.

| Variable | Perceived Stress | Salivary Cortisol | Hair Cortisol | Age | BMI | Symptom Burden | Bodily Distress | Spatial Extent of Pain | Tender Points | Pain Severity | Pain Interference | Pain Duration |
| --- | --- | --- | --- | --- | --- | --- | --- | --- | --- | --- | --- | --- |
| Perceived Stress |  |  |  |  |  |  |  |  |  |  |  |  |
| Salivary Cortisol | .02 |  |  |  |  |  |  |  |  |  |  |  |
| Hair Cortisol | .06 | .05 |  |  |  |  |  |  |  |  |  |  |
| Age | -.09 | -.15 | .12 |  |  |  |  |  |  |  |  |  |
| BMI | .106 | -.13 | .00 | .14 |  |  |  |  |  |  |  |  |
| Symptom Burden | .64** | -.05 | .07 | .11 | .29** |  |  |  |  |  |  |  |
| Bodily Distress | .68** | -.07 | .06 | .10 | .24** | .81** |  |  |  |  |  |  |
| Spatial Extent of Pain | .51** | -.10 | .00 | .19* | .30** | .79** | .73** |  |  |  |  |  |
| Tender Points | .44** | -.15 | -.01 | .23** | .38** | .79** | .68** | .84** |  |  |  |  |
| Pain Severity | .63** | -.13 | .02 | .16 | .32** | .85** | .82** | .80** | .78** |  |  |  |
| Pain Interference | .68** | -.11 | .05 | .11 | .32** | .82** | .86** | .75** | .71** | .92** |  |  |
| Pain Duration | .35** | -.13 | .04 | .33** | .23** | .44** | .42** | .55** | .53** | .48** | .46** |  |
| Fibromyalgiansess | .58** | -.10 | .03 | .18* | .25** | .85** | .76** | .97** | .84** | .84** | .78** | .56** |

*Note.* \* $p < .05$ , \*\* $p < .01$ . Perceived Stress = Perceived Stress Scale (PSS), BMI = Body Mass Index, Symptom Burden = Somatic Symptom Scale (SSS-8), Bodily Distress = Somatic symptom Disorder (SSD12), Spatial Extent of Pain = Widespread Pain Index (WPI), Fibromyalgiansess = Polysymptomatic Distress Scale (PSD).

### Exploratory data analyses

Additional exploratory analyses were performed to investigate subgroups of individuals with FMS regarding the stress indicators. A correlation analysis was therefore performed to investigate the associations of the clinical outcomes within the FMS sample. In this exploratory approach, a negative significant correlation was found between age and salivary cortisol, indicating, that the older the sample, the lower the salivary cortisol levels (*r* = .25, *p* < .05). The correlation results can be found in Appendix B and C in the supplementary material.

Another exploratory approach was to investigate whether there are subgroups of high or low cortisol profiles in individuals suffering from FMS. The FMS group was therefore divided into six subgroups with (1) high or (2) low salivary cortisol levels, and (3) high or (4) low hair cortisol levels and another two groups, one characterized by (5) high salivary and high hair cortisol and the other one characterized by (6) low salivary and low hair cortisol. These analyses revealed significant negative associations of the low-salivary cortisol group (n = 31) between salivary cortisol and bodily distress (*r* = -.355, *p* = 0.049) and pain severity (*r* = -.42, *p* = .016), and negative significant associations of the high-hair cortisol group (n = 42) between hair cortisol and pain duration (*r* = -.323, *p* = .035). In the group with low salivary and low hair cortisol (n = 16), a negative association was found between pain severity and salivary cortisol (*r* = -.52, *p* = .03).

In addition, the Cortisol Awakening Response (CAR) and the Area under the Curve (AUC) were exploratorily investigated, as these are other common methods for the investigation of salivary cortisol [44]. Both parameters did not show any significant differences between the groups, neither CAR *F*(1,139) = 1.52, *p* = .217, η_p_^2^ = 0.010, nor AUC (*F*(1,129) = .71, *p* = .399, η_p_^2^ = 0.006).

## Discussion

The primary aim of this study was to explore potential differences in stress levels between individuals with FMS and pain-free controls across several stress dimensions, including both perceived stress and endocrine indicators of stress. Stress assessment included three different dimensions: perceived stress levels, and daily average salivary cortisol and hair cortisol concentrations as indicators of acute and chronic stress levels via the HPA-axis. This comprehensive approach allowed us to identify and compare stress indicators across a broader spectrum. The main findings of this study were that (1) individuals with FMS reported significantly higher levels of perceived stress compared to pain-free controls, while (2) the cortisol data did not show significant group differences. Furthermore, correlation analysis showed that (3) clinical symptoms were correlated closely with perceived stress but not with cortisol indicators. No significant associations were observed between the stress dimensions themselves.

The finding that perceived stress in the last month was higher in individuals with FMS compared to controls, while there is no evidence on difference on cortisol markers of stress, is partly in line with the literature. As several meta-analyses have shown in recent years the evidence on cortisol levels in FMS is still very inconsistent and controversial [7; 23; 29]. For example, Fischer et al. [14], found no significant differences in hair cortisol concentrations between individuals with somatic functional disorders, including FMS, and healthy controls. Further, similar to our data, they also found no association between self-reported stress and hair cortisol. In addition, Coppens et al. [10] found no significant differences in baseline salivary cortisol, but significant differences in perceived stress between individuals with FMS and healthy controls. However, these results are in contrast to other studies that reported positive associations between hair and salivary cortisol [45] or hair cortisol and perceived stress [31], although effects were weak. Research in this area presents challenges that can significantly impact study results. Studies on salivary cortisol use different designs, sample numbers, and assay methods, leading to heterogenous outcomes [44; 46]. Hair cortisol is seen as a more stable long-term for chronic stress [30] due to some advantages over salivary cortisol, but factors like hair washing frequency or physical activity can affect the results [1; 11; 16; 37]. It is unlikely that a single stress measure can fully capture the activity of the body’s stress response system, as there is a complex interplay between multiple biological systems.

Our results show no significant associations between cortisol indictors of acute and chronic stress levels and clinical symptoms. However, when interpreting these data, it should be borne in mind that our cortisol measures only represent the systemic cortisol response, particularly that of the HPA-axis. We did not examine factors such as the sympathoadreno-medullary (SAM) axis or the neuroimmunological stress response. Therefore, we cannot extend our results to indicators of stress system activity beyond HPA activity. However, the SAM-axis and the body’s inflammatory system and their interaction with the brain’s neuronal networks play an important role in coping with stress [12].

In this regard, the imbalance of threat and soothing systems theory of stress by Pinto et al. [41], as well as the generalized unsafety theory [8] offer interesting concepts. Here the interaction of bodily cues, impaired interoception, challenging social contexts and the potential amplification of these factors by acute and chronic stress are emphasized. This multifaceted perspective underscores the critical importance of addressing stress, both acute and chronic, in the assessment and treatment of fibromyalgia. Psychological factors consequently play a crucial role in the perception and handling of pain, and stress in turn is associated with such psychological features. Internal and external control, for example, have relevant influences on how patients may cope and handle their pain in everyday life and how they may respond to treatments [32–35]. It is important to recognize that the subjective experience of stress results from the interconnection of all these components. In parallel, clinical symptom burden is strongly influenced by daily experiences that are embedded in a neural network of the brain that includes emotional and evaluative aspects that may lead to a more sensitive response to stress and pain [41]. Thus, consideration of individual markers of HPA activity alone is not sufficient to describe the stress response in patients with FMS.

Even though no significant differences were found at the overall group level, this does not mean that the HPA-axis is completely uninvolved in the complex interplay of FMS pathophysiology. While the main results of our study suggest a dissociation of the examined indicators, clinical correlations for cortisol were found in exploratory subgroup analyses. Subgroups of individuals with FMS were identified based on their cortisol profiles, including high and low salivary or hair cortisol groups, and combinations of high and low cortisol on both measures. These subgroups showed different associations with clinical outcomes such as physical distress, pain severity, pain duration and pain interference.

While the exploratory nature of the findings limits interpretation, it could suggest that there may be an association with clinical symptoms, particularly in certain subgroups and in cases of extreme HPA-axis dysregulation. Further research should therefore focus on the development of clinical approaches targeting subjectively perceived stressors, together with a broad investigation of a wide range of biomarkers using of multi-omics approaches to determine phenotypes of the HPA-axis to investigate the biological network underlying FMS, to clarify and better understand the pathophysiology.

### Limitations

Some limitations need to be mentioned. Next to unequal samples sizes of our compared groups, sex was not equally distributed. However, the adjustment for sex did not reveal significant influences on the results. Further, hair cortisol was not controlled for influencing factors such as hair washing frequency, shampooing or hair coloring. Neither did we control for physical activity, nor for the menstrual cycle of the participants. The data were collected during the COVID-19 pandemic, which was a stressor that had effects on physical and mental health and human behavior. This may have affected the stress measures. Furthermore, the time frames of the stress indicators only partially overlapped. This limits the interpretation of the results and underlines the question whether the chosen indicators were sufficient to represent acute and chronic stress.In such cases, experience sampling methods may provide better approaches to collect data on perceived stress while collecting biological stress measures. These limitations may restrict the interpretation of findings, however, prescribed standards for the analysis and interpretations were applied.

## Conclusion

In our study of FMS individuals and pain-free controls, individuals with FMS reported significantly higher subjective stress levels, closely related to symptom severity. Importantly, a dissociation between perceived stress and cortisol indicators of stress was observed. We found no evidence linking FMS to HPA axis-related markers of acute and chronic stress levels like cortisol concentrations in saliva or hair. This underscores the need for nuanced clinical approaches that target perceived stress in individuals with FMS to improve symptom management and to reveal the complex relationship between stress perception and physiological stress responses.

## Data Availability

All data produced in the present study are available upon reasonable request to the authors

## Acknowledgments

The submitted manuscript does not contain information about medical device(s)/drug(s). There are no conflicts of interest. This work was supported by the Deutsche Forschungsgemeinschaft (DFG) within Collaborative Research Center 1158 on pain and by the Bundesministerium für Bildung und Forschung; PerPAIN consortium, FKZ:01EC1904A). No benefits in any form have been or will be received from a commercial party directly or indirectly related to the subject of this manuscript.

## Supplement

**Appendix A.**
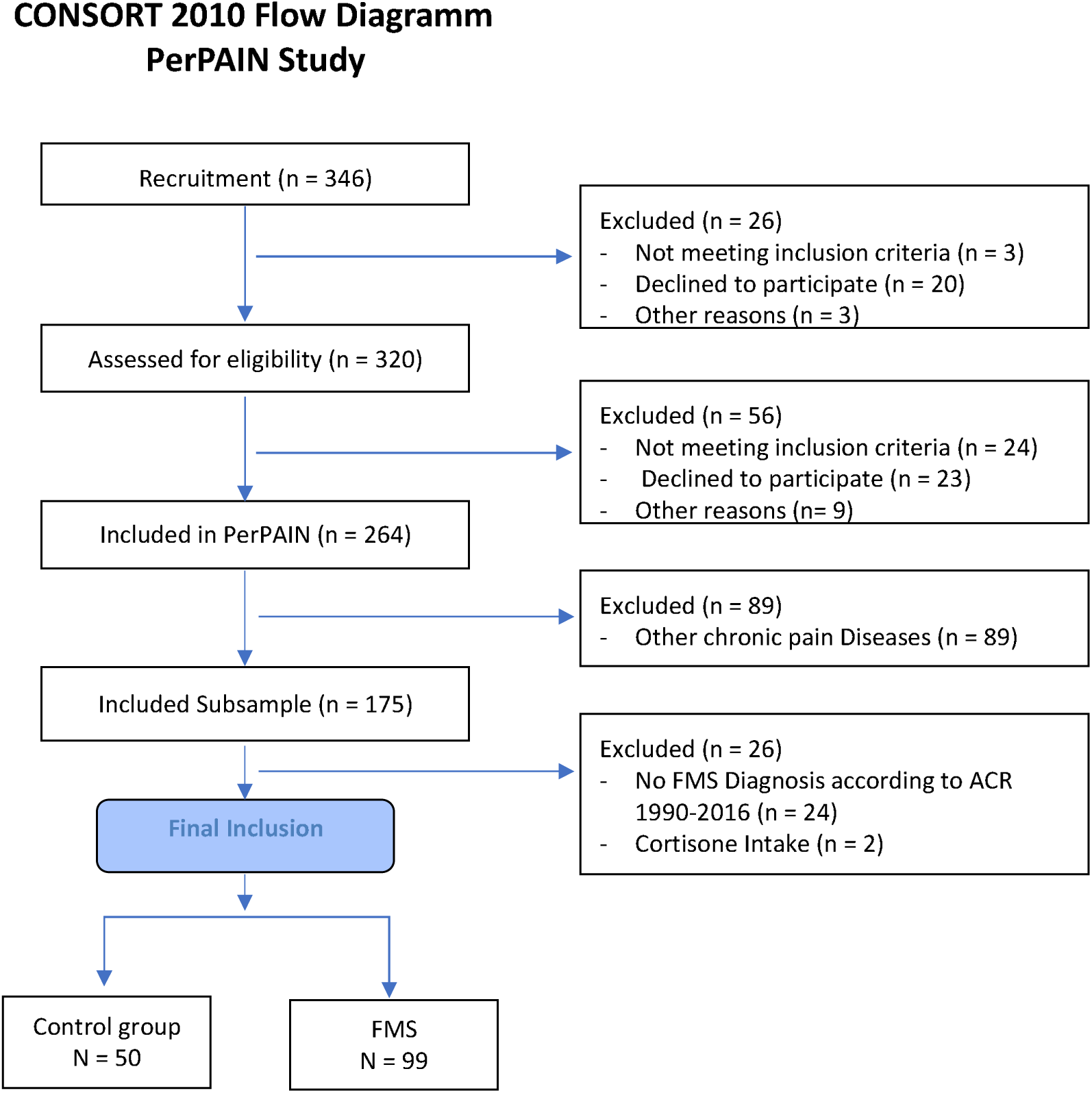
Flow Chart.

**Appendix B.** Correlation coefficients of stress indicators for the FMS sample N = 99.

| Variable | Perceived Stress | Salivary Cortisol | Hair Cortisol | Age | BMI | Symptom Burden | Bodily Distress | Spatial Extent of Pain | Tender Points | Pain Severity | Pain Interference | Pain Duration |
| --- | --- | --- | --- | --- | --- | --- | --- | --- | --- | --- | --- | --- |
| Perceived Stress |  |  |  |  |  |  |  |  |  |  |  |  |
| Salivary Cortisol | .13 |  |  |  |  |  |  |  |  |  |  |  |
| Hair Cortisol | .17 | .06 |  |  |  |  |  |  |  |  |  |  |
| Age | -.06 | -.25* | .01 |  |  |  |  |  |  |  |  |  |
| BMI | -.03 | -.14 | .07 | .10 |  |  |  |  |  |  |  |  |
| Symptom Burden | .41** | -.08 | .03 | .03 | .14 |  |  |  |  |  |  |  |
| Bodily Distress | .56** | -.06 | .03 | -.04 | .02 | .46** |  |  |  |  |  |  |
| Spatial Extent of Pain | .03 | -.13 | -.11 | .17 | .13 | .29** | .10 |  |  |  |  |  |
| Tender Points | -.02 | -.18 | -.09 | .32** | .29** | .44** | .13 | .43** |  |  |  |  |
| Pain Severity | .39** | -.19 | -.06 | .13 | .19 | .48** | .41** | .06 | .28** |  |  |  |
| Pain Interference | .55** | -.10 | .01 | .00 | .17 | .48** | .61** | .11 | .17 | .72** |  |  |
| Pain Duration | .05 | -.13 | .04 | .39** | .09 | -.03 | -.07 | .07 | .11 | -.05 | .00 |  |
| Fibromyalgianess | .25* | -.12 | -.08 | .10 | .02 | .50** | .18 | .82** | .44** | .22* | .23* | .09 |

Note. \* $p < .05$ , \*\* $p < .01$ .
Perceived Stress = Perceived Stress Scale (PSS), BMI = Body Mass Index, Symptom Burden = Somatic Symptom Scale (SSS-8), Bodily Distress = Somatic symptom Disorder (SSD12), Spatial Extent of Pain = Widespread Pain Index (WPI), Fibromyalgianess = Polysymptomatic Distress Scale (PSD)

**Appendix C.**
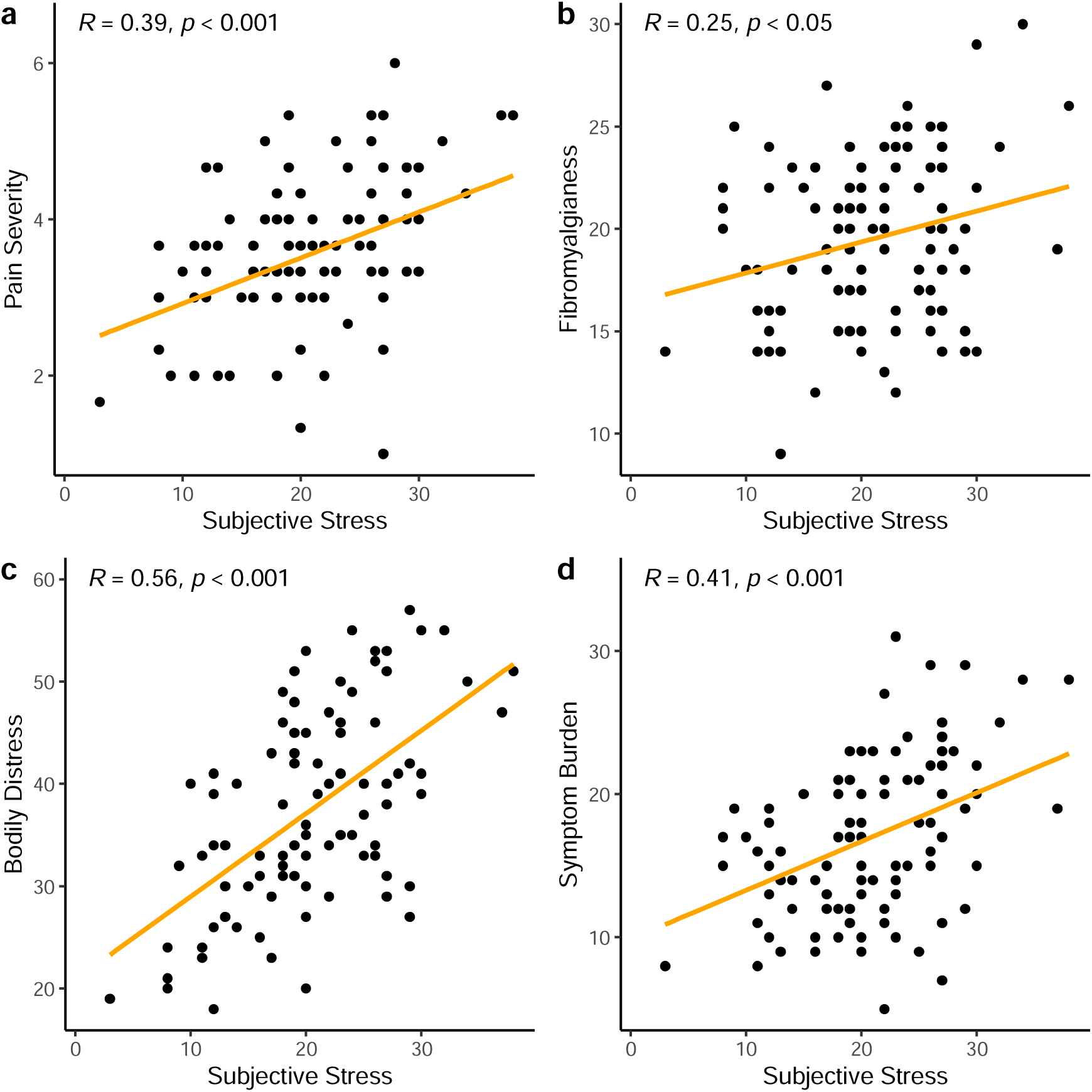
Correlation coefficients of stress indicators for the FMS sample N = 99. Scatterplots. Correlations between perceives Stress (PSS) with clinical outcomes (a) Pain Severity, (b) Fibromyalgianess = Polysymptomatic Distress Scale (PSD), (c) Bodily Distress = Somatic Distress Scale (SSD12), (d) Symptom Burden = Somatic Symtom Scale (SSS-8), for the FMS sample only (N = 99).

## Notes

### Competing Interest Statement

The authors have declared no competing interest.

### Clinical Protocols

https://archive.org/details/osf-registrations-g5du7-v1

### Funding Statement

The study was funded by the German Federal Ministry of Education and Research (PerPAIN consortium, FKZ:91EC1904A).

### Author Declarations

The Ethics Research Committee II of the Faculty of Medicine, University of Heidelberg (2020-579N), gave ethical approval for this work.

## References

[1] Abell JG, Stalder T, Ferrie JE, Shipley MJ, Kirschbaum C, Kivimäki M, Kumari M. Assessing cortisol from hair samples in a large observational cohort: The Whitehall II study. Psychoneuroendocrinology 2016;73:148–156.

[2] Adam EK, Quinn ME, Tavernier R, McQuillan MT, Dahlke KA, Gilbert KE. Diurnal cortisol slopes and mental and physical health outcomes: A systematic review and meta-analysis. Psychoneuroendocrinology 2017;83:25–41.

[3] Afari N, Ahumada SM, Wright LJ, Mostoufi S, Golnari G, Reis V, Cuneo JG. Psychological trauma and functional somatic syndromes: a systematic review and meta-analysis. Psychosomatic medicine 2014;76(1):2–11.

[4] Behrends J, Bischofberger J, Deutzmann R, Ehmke H, Frings S, Grissmer S, Hoth M, Kurtz A, Leipziger J, Müller F. Duale Reihe Physiologie [Dual Series Physiology]: Thieme, 2021.

[5] Beiner E, Baumeister D, Buhai D, Löffler M, Löffler A, Schick A, Ader L, Eich W, Sirazitdinov A, Malone C. The PerPAIN trial: a pilot randomized controlled trial of personalized treatment allocation for chronic musculoskeletal pain—a protocol. Pilot and Feasibility Studies 2022;8(1):1–12.

[6] Beiner E, Kleinke K, Tesarz J. Perceived, endocrine acute and chronic stress indicators in Fibromyalgia Syndrome. Publication in preparation., 2024.

[7] Beiner E, Lucas V, Reichert J, Buhai D-V, Jesinghaus M, Vock S, Drusko A, Baumeister D, Eich W, Friederich H-C. Stress biomarkers in individuals with fibromyalgia syndrome: a systematic review with meta-analysis. PAIN 2023;164(7):1416–1427.

[8] Brosschot JF, Verkuil B, Thayer JF. Generalized unsafety theory of stress: Unsafe environments and conditions, and the default stress response. International journal of environmental research and public health 2018;15(3).

[9] Cohen S, Kamarck T, Mermelstein R. A global measure of perceived stress. Journal of health and social behavior 1983;24(4):385–396.

[10] Coppens E, Kempke S, Van Wambeke P, Claes S, Morlion B, Luyten P, Van Oudenhove L. Cortisol and subjective stress responses to acute psychosocial stress in fibromyalgia patients and control participants. Psychosomatic Medicine 2018;80(3):317–326.

[11] Dettenborn L, Tietze A, Kirschbaum C, Stalder T. The assessment of cortisol in human hair: associations with sociodemographic variables and potential confounders. Stress 2012;15(6):578–588.

[12] Dorsey A, Scherer E, Eckhoff R, Furberg R. Measurement of human stress: a multidimensional approach. RTI Press 2022.

[13] Enders CK. Applied missing data analysis: Guilford Publications, 2022.

[14] Fischer S, Skoluda N, Ali N, Nater UM, Mewes R. Hair cortisol levels in women with medically unexplained symptoms. Journal of Psychiatric Research 2022;146:77–82.

[15] Gao W, Stalder T, Foley P, Rauh M, Deng H, Kirschbaum C. Quantitative analysis of steroid hormones in human hair using a column-switching LC–APCI–MS/MS assay. Journal of Chromatography B 2013;928:1–8.

[16] Gerber M, Brand S, Lindwall M, Elliot C, Kalak N, Herrmann C, Pühse U, Jonsdottir IH. Concerns regarding hair cortisol as a biomarker of chronic stress in exercise and sport science. Journal of sports science & medicine 2012;11(4):571–581.

[17] Gierk B, Kohlmann S, Kroenke K, Spangenberg L, Zenger M, Brähler E, Löwe B. The somatic symptom scale–8 (SSS-8): a brief measure of somatic symptom burden. JAMA internal medicine 2014;174(3):399–407.

[18] Greff MJ, Levine JM, Abuzgaia AM, Elzagallaai AA, Rieder MJ, van Uum SH. Hair cortisol analysis: An update on methodological considerations and clinical applications. Clinical biochemistry 2019;63:1–9.

[19] Harris PA, Taylor R, Thielke R, Payne J, Gonzalez N, Conde JG. Research electronic data capture (REDCap)—a metadata-driven methodology and workflow process for providing translational research informatics support. Journal of biomedical informatics 2009;42(2):377–381.

[20] Heim C, Nater UM, Maloney E, Boneva R, Jones JF, Reeves WC. Childhood trauma and risk for chronic fatigue syndrome: association with neuroendocrine dysfunction. Archives of general psychiatry 2009;66(1):72–80.

[21] Hemmerich W. StatistikGuru: Bonferroni-Holm-Korrektur [StatistikGuru: Bonferroni-Holm correction], 2020.

[22] Holm S. A simple sequentially rejective multiple test procedure. Scandinavian journal of statistics 1979;6(2):65–70.

[23] Illescas-Montes R, Costela-Ruiz VJ, Melguizo-Rodríguez L, Luna-Bertos D, Ruiz C, Ramos-Torrecillas J. Application of Salivary Biomarkers in the Diagnosis of Fibromyalgia. Diagnostics 2021;11(1):63.

[24] Kaleycheva N, Cullen AE, Evans R, Harris T, Nicholson T, Chalder T. The role of lifetime stressors in adult fibromyalgia: systematic review and meta-analysis of case-control studies. Psychological medicine 2021;51(2):177–193.

[25] Kerns RD, Turk DC, Rudy TE. The west haven-yale multidimensional pain inventory (WHYMPI). Pain 1985;23(4):345–356.

[26] Kleinke K, Reinecke J, Salfrán D, Spiess M. Applied multiple imputation: Springer, 2020. [27] Koğar E, Koğar H. A systematic review and meta-analytic confirmatory factor analysis of the perceived stress scale (PSS-10 and PSS-14). STRESS AND HEALTH 2023.

[28] Kozlov A, Kozlova M. Cortisol as stress marker. Human physiology 2014;40(2):224–236.

[29] Kumbhare D, Hassan S, Diep D, Duarte FC, Hung J, Damodara S, West DW, Selvaganapathy PR. Potential role of blood biomarkers in patients with fibromyalgia: a systematic review with meta-analysis. Pain 2022;163(7):1232–1253.

[30] Lee DY, Kim E, Choi MH. Technical and clinical aspects of cortisol as a biochemical marker of chronic stress. BMB reports 2015;48(4):209–216.

[31] Lynch R, Flores-Torres MH, Hinojosa G, Aspelund T, Hauksdóttir A, Kirschbaum C, Catzin-Kuhlmann A, Lajous M, Valdimarsdottir U. Perceived stress and hair cortisol concentration in a study of Mexican and Icelandic women. PLOS Global Public Health 2022;2(8):e0000571.

[32] Malin K, Littlejohn GO. Personality and fibromyalgia syndrome. The Open Rheumatology Journal 2012;6:273–285.

[33] Malin K, Littlejohn GO. Psychological control is a key modulator of fibromyalgia symptoms and comorbidities. Journal of Pain Research 2012;5:463–471.

[34] Malin K, Littlejohn GO. Stress modulates key psychological processes and characteristic symptoms in females with fibromyalgia. Clinical Experimental Rheumatology 2013;31(6, Suppl 79):64–71.

[35] Malin K, Littlejohn GO. Psychological factors mediate key symptoms of fibromyalgia through their influence on stress. Clinical rheumatology 2016;35(9):2353–2357.

[36] Marques AP, Santo AdSdE, Berssaneti AA, Matsutani LA, Yuan SLK. Prevalence of fibromyalgia: literature review update. Revista brasileira de reumatologia 2017;57:356–363.

[37] McEwen BS. What is the confusion with cortisol? Chronic Stress 2019;3:1–3.

[38] Nicholas M, Vlaeyen JW, Rief W, Barke A, Aziz Q, Benoliel R, Cohen M, Evers S, Giamberardino MA, Goebel A. The IASP classification of chronic pain for ICD-11: chronic primary pain. Pain 2019;160(1):28–37.

[39] Noushad S, Ahmed S, Ansari B, Mustafa U-H, Saleem Y, Hazrat H. Physiological biomarkers of chronic stress: A systematic review. International Journal of Health Sciences 2021;15(5):46–59.

[40] Pape H, Kurtz A, Silbernagl S. Physiologie: Stuttgart New York: Thieme Georg Verlag, 2018.

[41] Pinto AM, Geenen R, Wager TD, Lumley MA, Häuser W, Kosek E, Ablin JN, Amris K, Branco J, Buskila D. Emotion regulation and the salience network: a hypothetical integrative model of fibromyalgia. Nature Reviews Rheumatology 2023;19(1):44–60.

[42] R Core Team. R: A Language and Environment for Statistical Computing (Version 4.1.2) [Computer Software]: R Foundation for Statistical Computing, 2022.

[43] Russell E, Koren G, Rieder M, Van Uum S. Hair cortisol as a biological marker of chronic stress: current status, future directions and unanswered questions. Psychoneuroendocrinology 2012;37(5):589–601.

[44] Stalder T, Kirschbaum C, Kudielka BM, Adam EK, Pruessner JC, Wüst S, Dockray S, Smyth N, Evans P, Hellhammer DH. Assessment of the cortisol awakening response: Expert consensus guidelines. Psychoneuroendocrinology 2016;63:414–432.

[45] Stalder T, Steudte-Schmiedgen S, Alexander N, Klucken T, Vater A, Wichmann S, Kirschbaum C, Miller R. Stress-related and basic determinants of hair cortisol in humans: A meta-analysis. Psychoneuroendocrinology 2017;77:261–274.

[46] Stoffel M, Neubauer AB, Ditzen B. How to assess and interpret everyday life salivary cortisol measures: A tutorial on practical and statistical considerations. Psychoneuroendocrinology 2021;133.

[47] Tak LM, Cleare AJ, Ormel J, Manoharan A, Kok IC, Wessely S, Rosmalen JG. Meta-analysis and meta-regression of hypothalamic-pituitary-adrenal axis activity in functional somatic disorders. Biological psychology 2011;87(2):183–194.

[48] Toussaint A, Riedl B, Kehrer S, Schneider A, Löwe B, Linde K. Validity of the Somatic Symptom Disorder–b Criteria Scale (ssd-12) in primary care. Family Practice 2018;35(3):342–347.

[49] Van Buuren S, Groothuis-Oudshoorn K. mice: Multivariate Imputation by Chained Equations in R. Journal of Statistical Software 2011;45(3):1–67.

[50] Van Houdenhove B, Egle U, Luyten P. The role of life stress in fibromyalgia. Current rheumatology reports 2005;7(5):365–370.

[51] Van Houdenhove B, Egle UT. Fibromyalgia: A stress disorder? Psychotherapy and psychosomatics 2004;73(5):267–275.

[52] Wolfe F, Butler SH, Fitzcharles M, Häuser W, Katz RL, Mease PJ, Rasker JJ, Russell AS, Russell IJ, Walitt B. Revised chronic widespread pain criteria: development from and integration with fibromyalgia criteria. Scandinavian journal of pain 2019;20(1):77–86.

[53] Wolfe F, Clauw DJ, Fitzcharles M-A, Goldenberg DL, Häuser W, Katz RL, Mease PJ, Russell AS, Russell IJ, Walitt B. 2016 Revisions to the 2010/2011 fibromyalgia diagnostic criteria. Seminars in arthritis and rheumatism 2016;46(3):319–329.

[54] Wolfe F, Clauw DJ, Fitzcharles M-A, Goldenberg DL, Häuser W, Katz RS, Mease P, Russell AS, Russell IJ, Winfield JB. Fibromyalgia criteria and severity scales for clinical and epidemiological studies: a modification of the ACR Preliminary Diagnostic Criteria for Fibromyalgia. The Journal of rheumatology 2011;38(6):1113–1122.

[55] Wolfe F, Clauw DJ, Fitzcharles MA, Goldenberg DL, Katz RS, Mease P, Russell AS, Russell IJ, Winfield JB, Yunus MB. The American College of Rheumatology preliminary diagnostic criteria for fibromyalgia and measurement of symptom severity. Arthritis care & research 2010;62(5):600–610.

[56] Wolfe F, Smythe HA, Yunus MB, Bennett RM, Bombardier C, Goldenberg DL, Tugwell P, Campbell SM, Abeles M, Clark P. The American College of Rheumatology 1990 criteria for the classification of fibromyalgia. Arthritis & Rheumatism: Official Journal of the American College of Rheumatology 1990;33(2):160–172.

[57] Wolfe F, Walitt B, Perrot S, Rasker JJ, Häuser W. Fibromyalgia diagnosis and biased assessment: Sex, prevalence and bias. PloS one 2018;13(9):e0203755.

[58] Wolfe F, Walitt BT, Rasker JJ, Katz RS, Häuser W. The use of polysymptomatic distress categories in the evaluation of fibromyalgia (FM) and FM severity. The Journal of rheumatology 2015;42(8):1494–1501.

